# A clinical MALDI-ToF Mass spectrometry assay for SARS-CoV-2: Rational design and multi-disciplinary team work

**DOI:** 10.1101/2020.08.22.20176669

**Authors:** Ray K Iles, Raminta Zmuidinaite, Jason K Iles, George Carnell, Alex Sampson, Jonathan L Heeney

**Affiliations:** MAPSciences The iLAB, Stannard Way, Bedford, UK MK44 3RZ; Laboratory of Viral Zoonotics, Department of Veterinary Medicine, Cambridge University, Madingley Road, Cambridge, UK CB3 0ES; NISAD, Medicon Village, Lund, Sweden

**Keywords:** COVID-19, SARS-CoV-2, screening diagnostic test, MALDI-ToF MS

## Abstract

The COVID-19 pandemic caused by the SARS-CoV-2 Coronavirus has stretched national testing capacities to breaking points in almost all countries of the world. The need to rapidly screen vast numbers of a country’s population in order to control the spread of the infection is paramount. However, the logistical requirement for reagent supply (and associated cost) of RT-PCR based testing (the current front-line test) have been hugely problematic. Mass spectrometry-based methods using swab and gargle samples have been reported with promise, but have not approached the task from a systematic analysis of the entire diagnostic process. Here, the pipeline from sample processing, the biological characteristics of the pathogen in human biofluid, the downstream bio- and physical-chemistry and the all-important data processing with clinical interpretation and reporting, are carefully compiled into a single high throughput and reproducible rapid process.

Utilizing MALDI-ToF mass spectrometric detection to viral envelope glycoproteins in a systems biology – multidisciplinary team approach, we have achieved a multifaceted clinical MALDI ToF MS screening test, primarily (but not limited to) SARS-CoV-2, with direct applicable to other future epidemics/pandemics that may arise.

The clinical information generated not only includes SARS-CoV-2 Coronavirus detection – (Spike protein fragments S1, S2b, S2a peaks), but other respiratory viral infections detected as well as an assessment of generalised oral upper respiratory immune response (elevated total Ig light chain peak) and a measure of the viral immune response (elevated intensity of IgA heavy chain peak).

The advantages of the method include; 1) ease of sampling, 2) speed of analysis, and much reduced cost of testing. These features reveal the diagnostic utility of MALDI-ToF mass spectrometry as a powerful and economically-attractive global solution.

**Abstract graphic:** 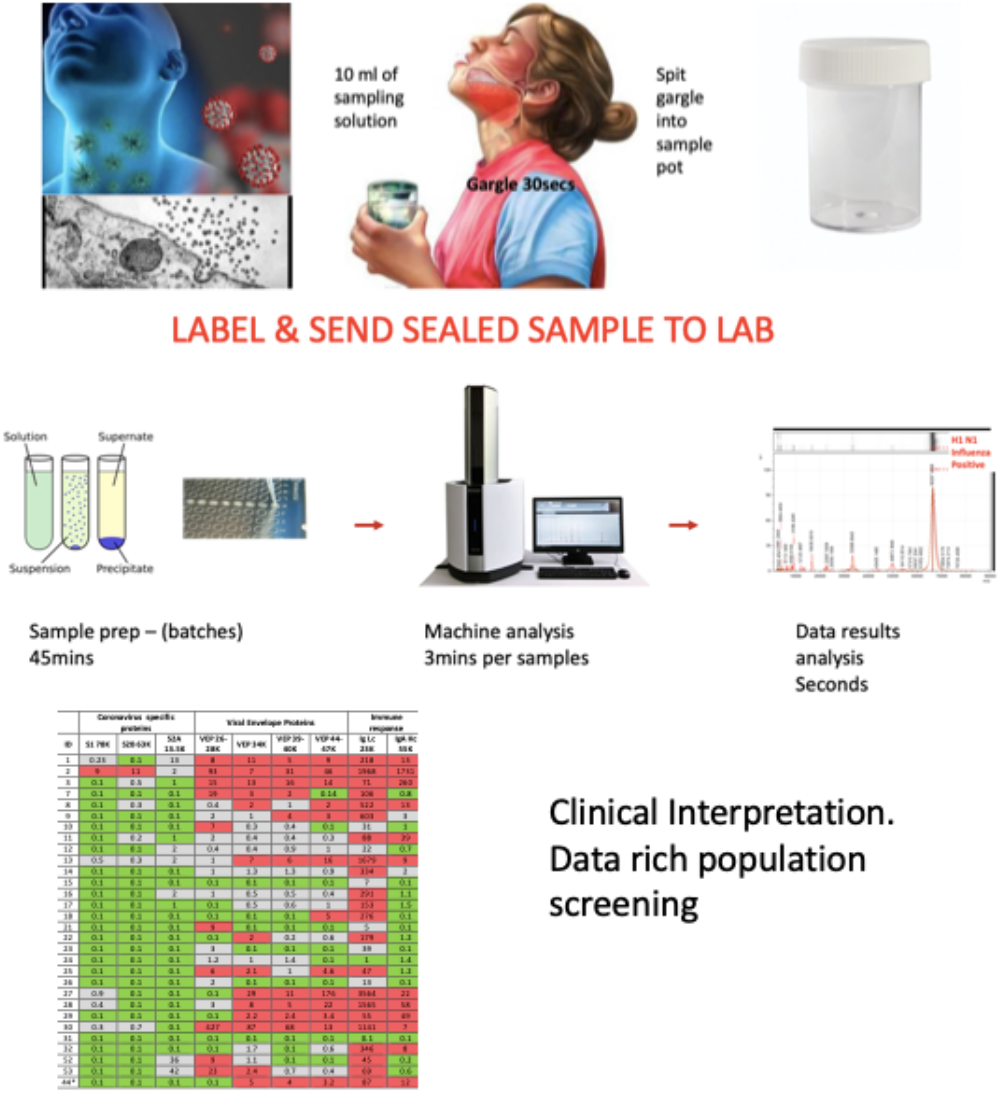

## Introduction

Mass screening of populations for infection in epidemics, but particularly pandemics, requires clinically effective (high sensitivity and selectivity) testing methods, requiring minimally-invasive sampling techniques, with fast turnaround (low hours) and a low cost of analysis (< 1 USD) a key feature in the context of large-scale screening of populations and the need for differential diagnosis of other circulating common cold pathogens.

Indeed, when the need for large-scale screening is global, logistical challenges such as the technical manpower, expertise, equipment and reagent availability and cost of the frontline test are less surmountable for most economically-challenged countries facing the pandemic, rendering control of the epidemic in many regions beyond reach.

Currently, RT-PCR testing is at the frontline for the identification of SARS-CoV-2 RNA. SARS-CoV-2 RNA is generally detectable in upper respiratory swabs and bronchoalveolar lavage (BAL) samples during the acute phase of infection. Positive results are indicative of the presence of SARS-CoV-2 RNA; while clinical plasma biomarkers such as D-dimer, CRP, IL-6 levels with patient history are necessary to determine the patient’s clinical COVID-19 status and level of therapeutic intervention.

The 2020 global pandemic caused by novel coronavirus SARS-CoV-2 pandemic has created an enormous challenge for health systems, clinical laboratories, public health intervention strategies, and community control measures worldwide. Testing limitations, including reagent shortages, remain a bottleneck in the battle to curtail COVID-19 spread in even the wealthiest countries [1,2]

The development of new matrix assisted laser desorption time of flight mass spectrometry (MALDIToF MS) diagnostics for SARS-CoV-2 detection is driven by the need for greater diagnostic capacity and alternative applications to complement standard PCR and antibody based diagnostics.

## Rational design

Preparative design in the development of any new assay system can circumvent many of the inevitable hurdles and dead ends that are encountered in early studies that have high promise, but have not considered detailed real-world characteristics. These include sampling errors, pathogen biology, biochemistry and physical limitations of the technology; not to mention due regard for bioinformatic rigor needed for classification and identification of robust and relevant markers. This has often been observed in the field of mass spectrometry, following sweeping assumptions and unrepresentative standards. However, rational multi-discipline based approaches to address these challenges, with step by step robust tests can result in significant diagnostic and socio-health economic returns [3].

To develop MALDI-ToF MS based diagnostics to specifically identify SARS-CoV-2, while distinguish this pathogen from other respiratory viruses, we had to overcome hurdles in the initial stages, looking beyond the traditional approachs by using established protocols routinely employed for bacterial identification [4].

## Viral Biological target for detection by MALDI-ToF

Enveloped viruses have the unique biological feature of utilizing the host cell membrane as the outer coating covering the core containing the RNA genome. The Coronaviruses have 4 key structural proteins and a large number of important functional proteins coded by their genome (Figure 1). Key amongst these viral envelope proteins are the Spike protein encompassing the receptor binding domain (RBD) and membrane fusion complex, domains that enable infection of target host cells. For the Coronaviruses these have been termed Spike or “S” proteins (see Figure 1a). Large trimeric complexes, these contain functional domains that undergo, what has been described as, tectonic conformational changes upon receptor binding and triggered exposure of membrane fusion peptide sequences [5,6]. These bring the viral envelope and target cell lipid bilayer membrane into close contact such that fusion can occur. Only a minor pore is sufficient for the viral RNA to enter and be expressed (figure 1b). Early Phase ORF expression takes control of the cell machinery, circumventing cellular defences such as ribonucleases and ubiquitination of viral proteins. Late phase ORF expression makes more viral envelope proteins, nucleoprotein and Spike complex transcribed and post-translationally modified by the host cells endoplasmic reticulum and Golgi apparatus. Together with the viral RNA genome, these are packaged into membrane exosomes to form enveloped virions bristling with spike protein complexes (see figure 1b and 1c).

**Figure 1:**
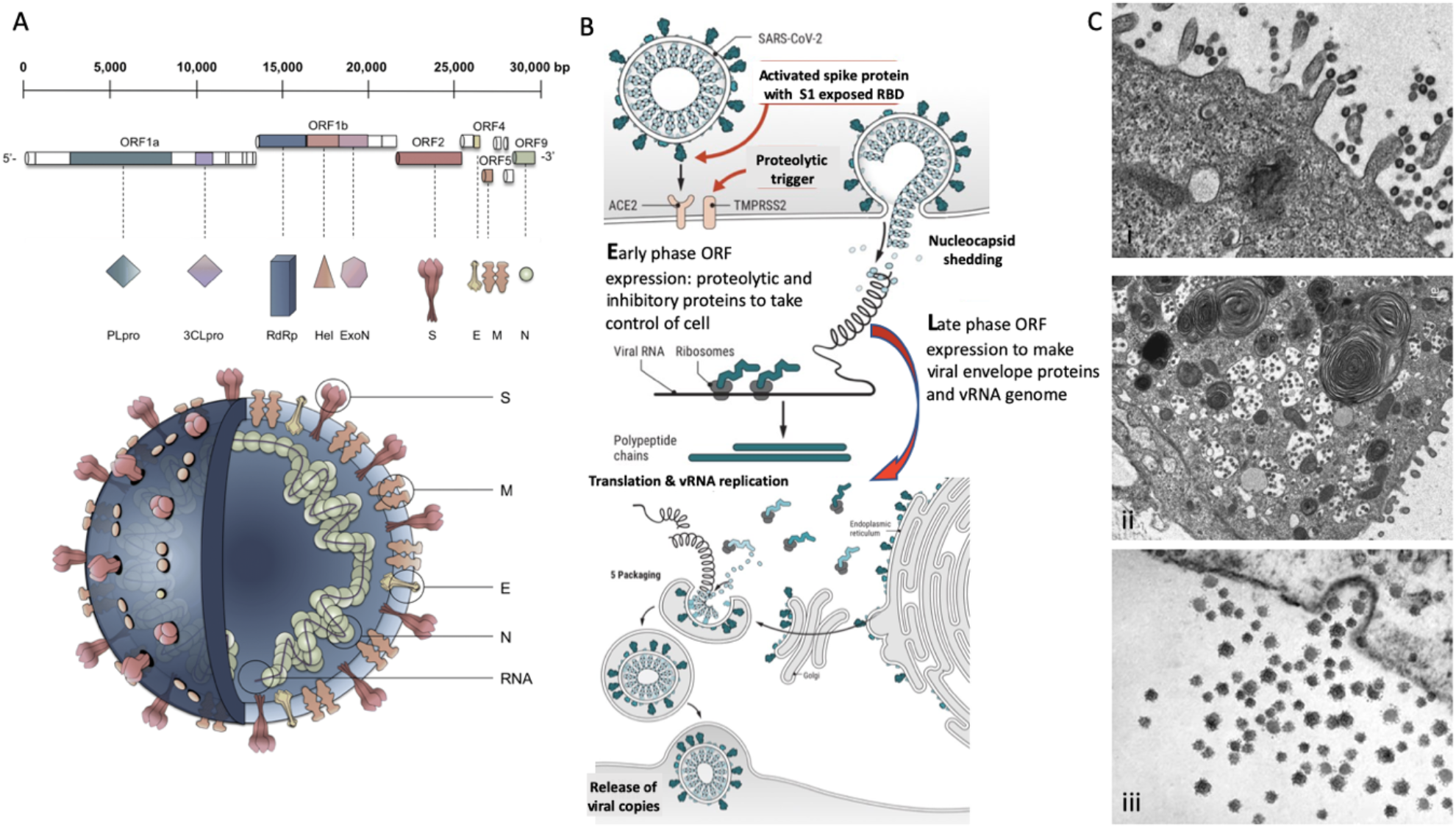
Molecular cellular pathology of SARS-CoV-2 infection: Panel A) Schematic Illustration of SARS-CoV-2 viral RNA genome in relation to order of expression and functional incorporation with the Virion particle. The genome comprises a 5ʹ-untranslated region (5ʹ-UTR), open-reading frames (ORFs) 1a and 1b encoding nonstructural proteins 3-chymotrypsin-like protease (3CLpro), papain-like protease (PLpro), helicase (Hel), and RNA-dependent RNA polymerase (RdRp) as well as accessory proteins, and the structural S protein(S), E protein (E), M protein (M), and Nucleocapsid protein (N) in ORFs 2,4,5 and 9. Panel B). Schematic of virion fusion mediated by Spike protein attachment to the ACE2 receptor and accessory cleavage of the Spike protein quaternary complex, resulting in fusion peptide exposure and binding to target cell plasma membrane. Focal, host cell and viral envelope lipid bilayer membrane fusion and viral RNA ingress of the target host cell. Shedding of Nucleocapsid and early ORF expression to take control of the cell and inactivate internal anti-viral proteins (e.g. Ubiquitin and RNase). Late ORF expression: replication of viral RNA genome and expression of, via post-translational processing mechanisms of the endoplasmic reticulum and Golgi apparatus (e.g. glycosylation), membrane embedded proteins that are responsible for infectivity including the Spike protein complex. Copies of SARS-CoV-2 vRNA are packaged into lipid bilayer membrane envelopes containing multiple copies of S, E, M and N, glyco- and phospho-proteins. Inclusion transport vesicle, packed with enveloped virions, fuses with host cell membrane releasing infective SARS-CoV-2 virion. Panel C). Electro micrographs of multiple virions attacking the cell membrane (i), viral genome expression subsuming all functions of the cell to form thousands of virion copies within inclusion vesicles (ii) and finally release of multiple virions by fusion of inclusion packaging vesicles with the cell surface plasma membrane of the infected host cell (iii).

## Optimisation of MALDI-ToF matrix for the identification of SARS-CoV-2 and other envelope virus embedded membrane proteins

In MALDI-ToF mass spectrometry for bacterial identification, the measured signals represent intracellular proteins, mostly highly abundant ribosomal proteins. The mass range usually measured ranges from 2,000 to 20,000 Th; the proteins revealed are largely not glycosylated, although they may be post-translationally modified in other ways such as phosphorylation [7]. The most appropriate robust and widely used matrix for ionisation of such unique or characteristic microbial intracellular proteins is *alpha*-cyano-4-hydroxycinnamic acid (CHCA) [8].

In consideration of a MALDI-ToF MS test for virus detection, this mass range is unsuitably low; significant differences in virion particles include the absence of intracellular “housekeeping” proteins. The majority of the unique features of viral proteins observed in MALDI are much larger (10,000 –300,000 Th),. Furthermore, many of the virus specific proteins are heavily glycosylated [9] and posttranslationally modified. This mass range is outside the effective limits for CHCA; other matrices have been shown to be more suited for the MALDI detection of large and glycosylated proteins [10].

Our method development investigated alternative matrices (data not shown);sinapinic acid (SA) (3-(4-hydroxy-3,5-dimethoxyphenyl)prop-2-enoic acid) proved to be the most consistent and versatile and was used in all subsequently developed protocols.

## Sampling, sample handling

Currently, sampling for COViD-19 screening in frontline testing is via nasopharyngeal and or oropharyngeal swabs, often an unpleasant experience for the individual being tested and a potential reason for poor compliance within regular testing regimes, particularly amongst children [11].

Once taken the swab sample has to be made safe for handling by medical, transport and laboratory staff. Current methods inactivate virus by destroying proteins and the viral envelope membrane, preserving nucleic acid. Generally, nasal-pharyngeal swabs are either heat inactivated, disrupting the protein and viral envelope membrane, or stored in a transport media containing SDS and or Triton which destroys the viral envelope of the virus, but is detrimental to mass spectrometric detection methods [12].

For effective MALDI-ToF MS COVID-19 testing, based on detection and quantification of viral proteins, clinical sample deactivation has to largely do the opposite, i.e. preserve viral membrane and proteins but destroy the functional nucleic acid. This is conveniently achieved by the UV-C irradiation of sample tubes, for as little as 15mins to ensure safe handling in the laboratory and by transport systems [13].

Consequently studies where swab samples have been split for simultaneous analysis by RT PCR detection systems of SARS-CoV-2 RNA and by MALDI-ToF mass spectrometry for viral proteins, are compromised [4].

As an alternative to swabs, saliva and/or gargle is an acceptable, and more easily obtained sample means of collection; 10mls of water was found to be the optimum collection volume media for gargle saliva testing (data not shown).

## Enrichment of viral proteins within the analyte sample

Whether viral culture media, saliva/gargle, serum or urine, an enrichment of the target (e.g. viral Envelope proteins), by the removal of unwanted entities is often necessary to improve mass spectrometric detection. he presence of abundant host molecules (such as albumin) can not only mask key, less abundant target proteins, but also suppress the ionisation of other low abundance proteins, even if they are detected at a distant m/z. Not only does enrichment, and clean-up improve the detection of lower abundance signals, but also improves shot-to-shot and sample-to-sample reproducibility [14,15].

In this work, the sample clean-up exploited the biophysical size difference of several orders of magnitude between virion particle and most components of acellular fractions of human biological fluids, allowing the convenient separation of them by precipitation. Separation by ultracentrifugation was ruled out as impractical for routine implementation, as was selective precipitation by salt addition or PEG, due to their deleterious impact on MS analysis. Acetone precipitation, using ice-cold addition, was found to be the most effective means of reproducible, facile selective precipitation for MS analysis. Other assisted precipitation methods such salting out resulted in samples being contaminated with high concentration of ions that suppressed target protein matrix assisted ionisation; whilst Polyethylene Glycol (PEG) assisted precipitation resulted in multiple interfering polymer peaks in the resultant MS spectra (data not shown).

The optimised methodology for efficiency, speed a simplicity consisted of a 1 to 1 addition of ice cold acetone, spun in a refrigerated bench top centrifuge at 16,000g for 30mins. The supernatant (containing unwanted background proteins) is discarded and the pellet resuspended in a membrane dissolution and viral envelope protein solubilisation buffer (figure 2). Much higher ratios of acetone and lower temperatures are commonly used but many more moderate to small proteins coprecipitate as a result and interfere with the final diagnostic mass spectra [16].

**Figure 2:**
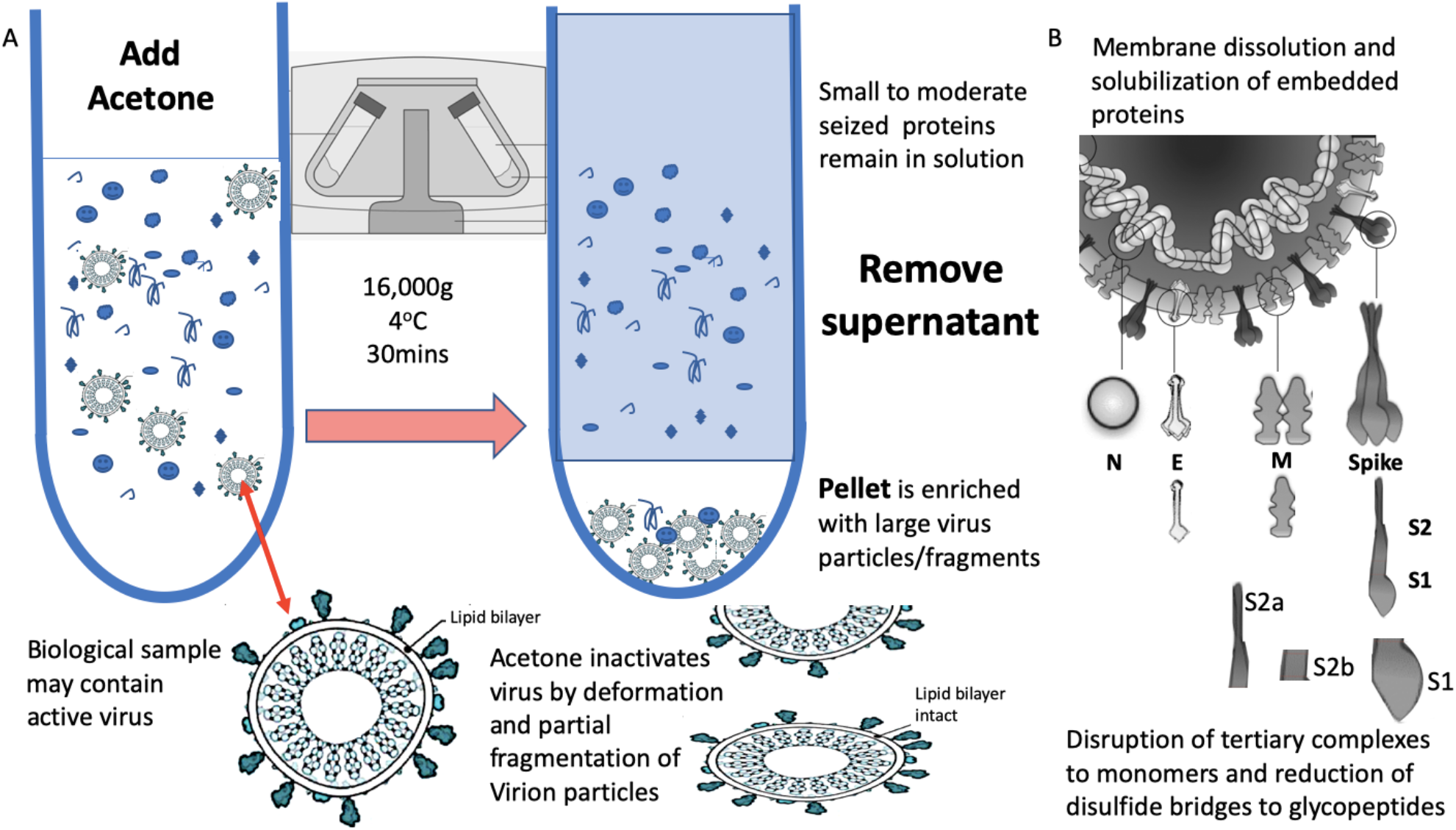
Process of virion enrichment within a biological sample and extraction/solubilisation of Spike (S) and other viral envelope protein for MALDI ToF MS analysis. Panel A: Biological samples containing virus are mixed 1 to 1 v/v with ice cold acetone and centrifuged at 16000g for 30 minutes at 4°C. The supernatant containing smaller non-precipitated solutes is discarded. The pellet is enriched with viral particles and kept for analysis. Acetone treatment inactivates envelope virus by deformation and partial fragmentation of the viral envelope and embedded protein structures. Panel B: The pellet is resuspended in 10 to 100ul of MALDI –ToF Mass spectrometry compatible dissolution and solubilisation buffer. Termed LBSD-X this buffer does not suppress ionisation and contains a detergent at a concentration optimised to release viral envelope embedded proteins together with non embedded viral proteins. It also contains Dithiothreitol (DTT) in order to further reduce disulphide bonds so that quaternary and tertial structures are fully disrupted and monomers, polypeptide chains and glyco-polypeptides are liberated for detailed mass analysis.

Acetone is an ideal organic solvent for the purpose of virion enrichment for mass spectrometry as not only does its weaker hydrogen and dipole bonding destabilise the hydration cloud surrounding large molecules, leading to precipitation, but it conveniently evaporates at room temperature without leaving a residue. Other organic solvents, including ethanol, similarly effect a precipitation but leave a residue [17]. This may account for acetone precipitation/evaporation having no detrimental effect on subsequent MALDI-ToF MS analysis. In addition, acetone denatures the viral envelope such that any virus is rendered biologically inert, but without completely destroying its structure. (Figure 2a) [18]. This is not the case for salting, PEG and even methanol co-precipitation enrichment of virion particles, where they have detrimental effects on ionisation & spectra, and have been shown to leave the virus biologically active with increased PFU potencies within the precipitation [19,20,21].

## Disruption of Viral Envelope and solubilisation of Viral proteins

Comparable to the analytical challenge Laemmli faced in his pioneering development of SDS –PAGE for viral protein analysis [22]; here, large complexes have to be disrupted and the viral proteins solubilised as free proteins to be seen by MALDI ToF Mass spectrometry (figure 2b). Unfortunately, detergents like SDS and Triton and CHAPS, although widely used in molecular biology, largely suppress ionization [23] and rapidly build up as fouling components on both the primary lens and other internal components of the mass spectrometer (supplemental figure A).

A series of novel detergents which avoided ion suppression and reduced lens fouling properties, were developed for solubilisation of hydrophobic and viral membrane bound proteins. The best performing formulation (data not shown), termed “LBSD-X” was adopted for all subsequent experiments.

In addition to solubilisation and ionization, the size of the target (glyco)proteins has to be considered with respect to the measurement range of the mass spectrometers. In our case best performance on the Shimadzu MALDI 8020 (Shimadzu-Kratos, Manchester, UK) was a maximum mass limit of 250,000 m/z. In more parallel’s to Laemmli’s work, reduction of disulphide bonds was necessary in order to completely disrupt the tertiary structure to component protein/peptide chains. The resultant protein-polypeptide chains and glyco-polypeptides chains, although large, being within the high resolution capacity of this bench top mass spectrometer. Dithiothreitol (DTT) is well tolerated by MALDI-ToF MS, does not suppress ionization and thus was incorporated in the matrix dissolution buffer formulation.

## Characterisation of Pseudo and live virus Spike envelope proteins by mass spectrometry

An important concern in test development for an infectious agent and particularly highly contagious, life threatening virus, is safety. The SARS spike S protein, a unique target as a marker of the infection, could be used and titrated into a biological sample in order to develop an analytical test with no risk to the development team [24]. However, although several recombinant SARS CoV-2 viral proteins are available these are not optimal as the starting material in a clinical mass spectrometry viral detection system. They fail to represent the true nature of the molecule as it exists in a real clinical analysis: The spike protein exist as a trimer of S proteins partially embedded within the viral envelope. It is extensively glycosylated and is also proteolytically cleaved to prime and activate it for receptor binding and then exposure of fusion peptide domains to allow ingress of the viral genome [5,25] (figure 1a and 1b). Recombinant proteins do not reflect this context and structural complexity, both of which complicate sample processing to achieve marker release and correct detection and quantification.

Pseudo-virus are constructs whereby virion-like particles, consisting of viral envelope exosomes, are made *in vitro*; lacking the viral genetic material as it is not replicated or incorporated (see figure 3) [26]. These are ideal non-infectious starting material upon which to develop a viral protein mass spectrometry tests. SARS2 Spike protein S genetic sequences were inserted into a target host human cell line, Hek293; so that fully post-translationally modified, membrane embedded viral proteins are expressed and tertiary structures are functionally intact. It was found that co-transfection with TMP322 was necessary for pseudo-virus vectors constructed to express the spike proteins as a functional ingress effector, i.e. fusion with a target cell (see figure 3). Through optimisation of the MALDI-ToF MS method we discovered that the S protein was cleaved into S1, S2b and S2a fragments although held together by disulphide bridges [27]. These fragments were then searched for in the cultures of live COVID-19 virus grown on Vero, African green monkey, cells and shed as functional virions into the culture media. Using the optimised protocol developed on pseudo-virus cultures the same spike fragments could be seen, S1 (79,000m/z) being the most prominent (see Figure 3).

**Figure 3.**
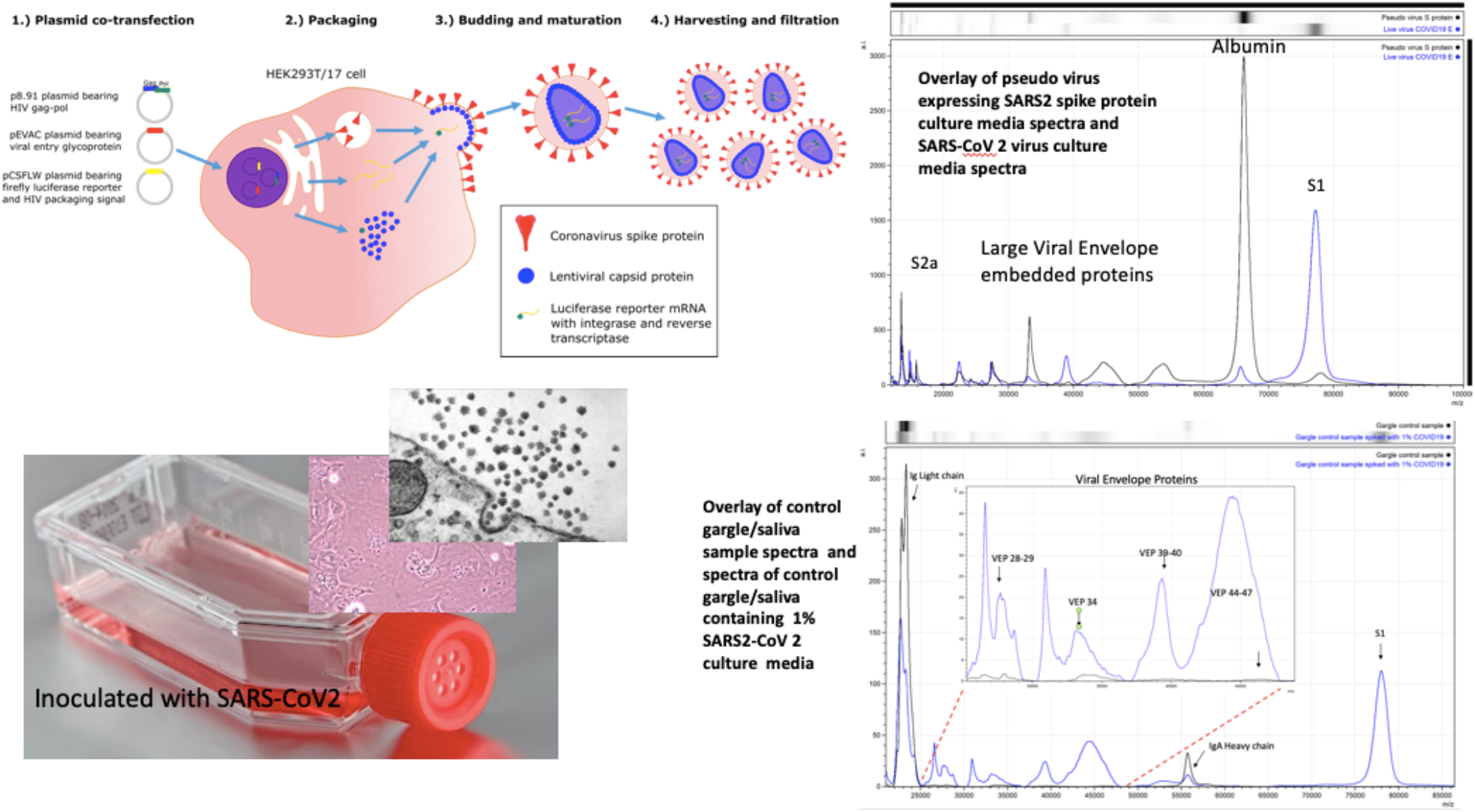
– Mass spectral profiles of Pseudo Virus expressing SARS-CoV-2 Spike S protein grown in culture, SARS-CoV-2 virus grown *in vitro* and mass spectra of gargle/saliva spiked with culture media from cells infected with SARS-CoV-2: S proteolytic fragments S1 and S2 were seen in all preparations and S2b only in serum free samples. Viral envelope proteins (VEPs) became more prevalent in live virus culture and Ig light chains and IgA heavy chain were additional peaks found in gargle/saliva samples. HEK293T/17 cells were seeded into 6 well cell-culture plates and co-transfected after 24 hours with p8.91 (gag-pol expression plasmid), pCSFLW (luciferase reporter plasmid), and a viral glycoprotein expression plasmid encoding the spike protein of either SARS-CoV-2. The transfection mixture was prepared in OptiMEM using FuGENE-HD as a transfection reagent; cell culture media was replaced prior to transfection. The cell supernatant was collected via syringe 48 hours after transfection and filtered through a 0.45 µm filter to harvest the lentiviral pseudo-type particles (26). Pseudo virus expressing post translationally modified and conformational function SARS S protein and live SARS-CoV-2 virus where grown in culture, under category 3 containment conditions, on Vero-Green Monkey cells. Virions present in the culture media were filtered (0.45µm), acetone precipitated-enriched as described and all viral envelope proteins, including S protein fragments, extracted & solubilized using extraction formulation LBSD-X buffer and detected as mass peaks by MALDI ToF MS.

Along with S protein fragments other viral component protein peaks were identified as characteristic of virions and were prominent in the viral cultures. These other Viral Envelope Proteins (VEPs) were characterised by their molecular mass, particularly a group of middle to high mass range broad peaks, indicative of variable glycosylation, which we have termed vep26–28k, vep34K, vep39–40K and vep45–47K.

## Identification of COVID-19 in salivary gargle samples

Having confirmed the specific and secondary target MS signals, indicative of corona virus, as S protein proteolytic fragments (primarily S1 and S2a) and VEPs as mass spectra peaks; the effect of the gargle/saliva sample milieu was examined for ‘matrix effect’ potential detrimental effects on the spectra. One striking example of a negative effect of natural biological sample matrix was clearly demonstrated by serum albumin, found in fetal calf serum (FCS) and used in supplementation of pseudo-virus and live virus cultures media. The mass peak of albumin obscuring detection of the similarly-sized S2b proteolytic fragment (figures 3 & 4).

**Figure 4.**
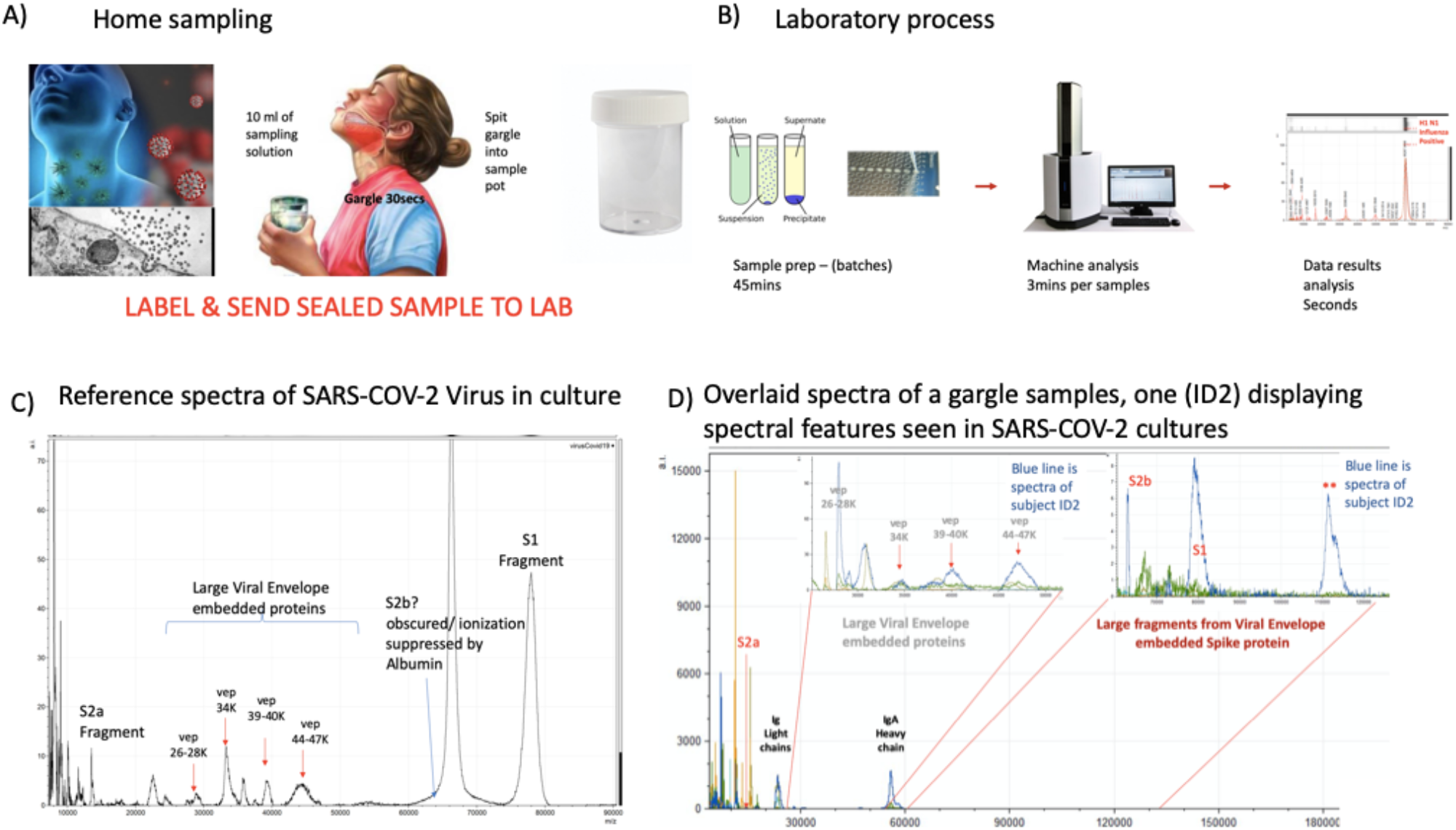
– Clinical diagnostic protocol of gargle saliva collection, laboratory processing and spectral data features: Panel A illustrates patient (home) sampling gargling 10ml of water and spitting that into a sample pot.Panel B illustrates rapid processing where received gargle sample is filtered through 1.2 and 0.45mm filters, 5ml is recovered and is acetone precipitated. The pellet is resuspended in viral envelope dissolution and protein solubilization buffer before being applied to MALDI ToF plates and analyzed (3mins per sample); output data being processed by appropriate software. Panel C Illustrates a reference spectra of SARS-CoV2 grown *in vitro* with spectral pattern of viral envelope proteins (VEPs) and fragments of Corona virus Spike protein labelled. Panel D illustrates overall spectra of 5 unscreened volunteer samples overlaid. The spectra from subject ID2 is indicated (blue line). Regions corresponding to mass ranges for VEPs and large characteristic fragments of the Spike protein are illustrated. These are illustrated in juxta position to gargle/salivary Immunoglobulin light chain and Ig A heavy chain peaks. Also to note that S2a fragment is surrounded by a host of high intensity/abundance proteins, the origins of which may be a mix of other co-enriched (bacterial) micro-organism from the oral cavity. An additional high molecular mass glycoprotein type peak was also seen at with a maxima at 112,000m/z (marked **) was only seen in this sample. Collection of volunteer gargle/saliva was by signed, informed consent and the study was approved by the IRB of MAPSciences and NISAD

Saliva is a size selective transudate of numerous serum proteins and organic biomolecules, often revealing these molecules at 10 to 100 fold lower concentrations than found in serum [28]. Albumin is fortunately not normally found at any significant levels in saliva and S2b should be evident in COVID-19 positive gargle/saliva samples. However, other large molecules such as amylase and IgA are actively secreted into saliva and IgA which has a mass of approximately 360,000 Th. The large proteins (such as IgA) are indeed co-precipitated in the acetone precipitation process described in this method, while the naturally free small to medium seized transudate and secreted proteins at lower masses, are removed in the supernatant. . Similarly, any other micro-organisms present in the sample along with any virus would also co –precipitated from the oral wash. To minimise potentially interfering effects, all gargle/saliva samples were sequentially prefiltered with 1.2 and 0.45µm syringe filters before acetone precipitation, retaining most large micro-organisms on the filter but allowing through virions and Immunoglobulins.

As a result of disulphide bond reduction, large molecular weight Immunoglobulins were visualised in the mass spectra of saliva samples, resolving as heavy and light chains (Figures 3 & 4). Their presence did not detract from spectral detection of the S1, S2b fragments or VEP’s. Despite being significant components.

However, by adding in live virus containing culture media to a control gargle saliva remained slightly compromised as a comparative testing model as BSA would always be present (Figures 3 & 4).

## Detection of COVID 19: Reproducibility, sensitivity and specificity

To estimate sensitivity and specificity saliva/gargle (n = 10) was spiked with spent Vero cell culture media producing SARS-CoV-2 at a concentration of 10^4^ to 10^5^ PFU/ml. These were subject to MALDIToF mass spectrometry and the peak heights measured for the peaks of interest. Two RT-PCR negative persons simultaneously provided gargle/saliva samples at the time of swab sampling. These confirmed PCR-negative gargle samples were analysed by MALDI-ToF mass spectrometry 40 times; the measured peak intensities of which acted as comparative controls to the viral spiked saliva/gargle. Viral envelope protein peak intensities were clearly elevated in the spiked samples (Figure 5). Gargle/saliva was collected from 35 volunteers who did not have simultaneous COVID-19 PCR determinations, although some suspected they had suffered from the infection but were recovered or recovering. The intensity of S protein fragments S1 (79,000m/z), S2b (62,000m/z) (can be obscured by albumin if present) and S2a (13,500m/z) were measured along with peaks masses thought to represent other viral envelope proteins (VEPs). The maximal intensity was measured for these broad glycoproteins peaks at 26,000–28,000m/z (vep26–28K), 34,000m/z (vep34K), 39,000 –40,000m/z (vep39–40K) and 45,000–47,000m/z (vep45–47K).

**Figure 5:**
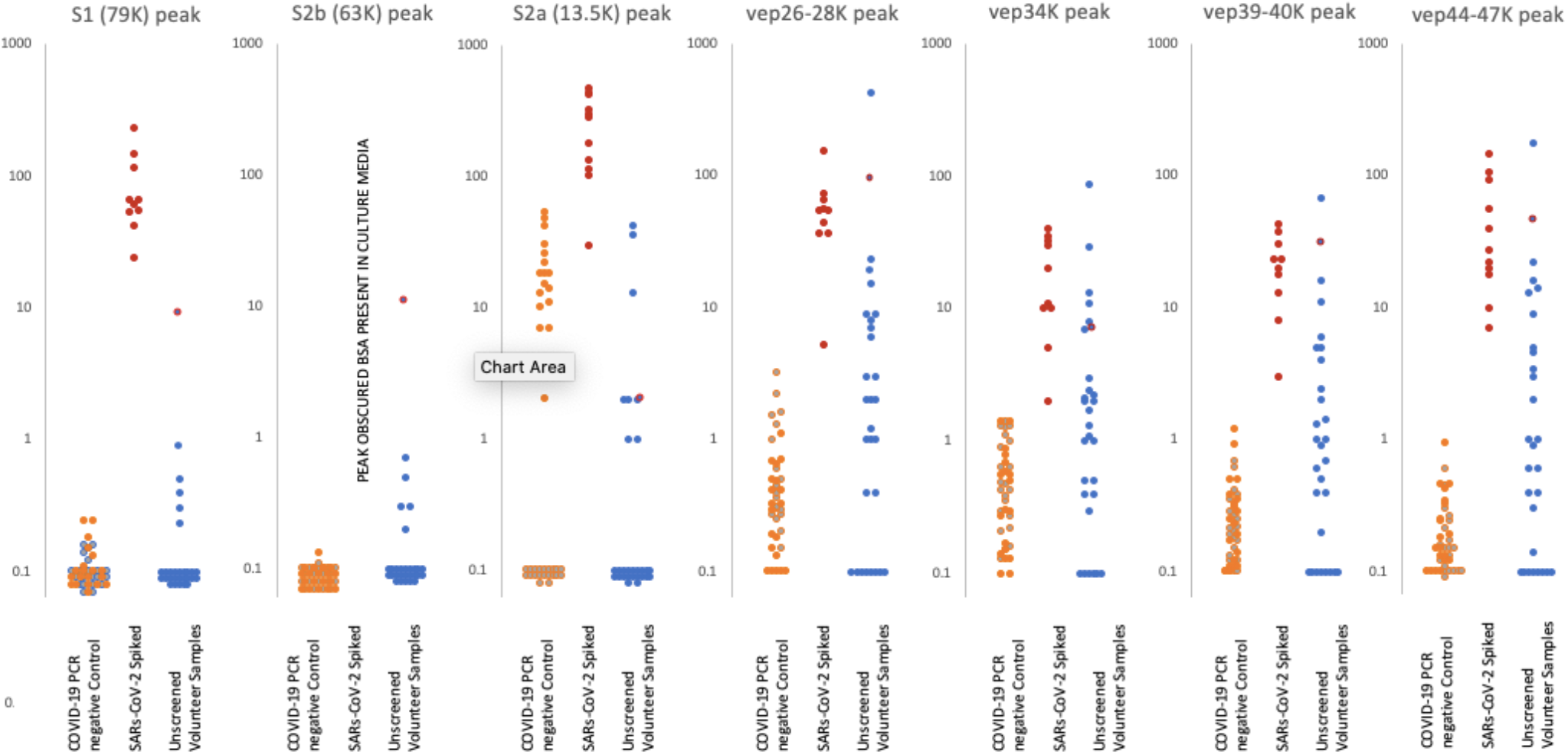
Levels of proteolytic S protein fragments and viral associated protein peaks in samples groups. Culture supernatant from Vero cells infected with Sars-Cov-2 were spiked into ten gargle/saliva samples at 1–10%. The viral load in these culture media was determined as at 10^4^ to 10^5^ PFU/ml. Gargle/saliva were collected from two individuals, with and without a cough, who were confirmed as RT-PCR Covid-19 negative at the time of gargle/saliva collection. These were analysed 20 times each. A series of 30 gargle/saliva was collected from volunteers, but COVID-19 PCR status at time of sampling was not known. These were analysed only once each. All samples were acetone precipitated, subjected to viral envelope protein solubilisation and analysed by MALDI-ToF mass spectrometry. Target peak intensities were then measured. Panel A) illustrates the levels of each analysed sample for Spike protein fragment S1 peak at 79,000m/z; panel B) Spike protein fragment S2b at 63,000m/z and panel C) Spike protein fragment S2a at 13,500m/z. The maximal intensity was also measured for broad glycoproteins peaks, considered to originate from other viral envelope proteins (VEPS), at 26,000–28,000m/z (vep26–28K – panel D), 34,000m/z (vep34K panel E), 39,000 –40,000m/z (vep39–40K – panel F) and 45,000 –47,000m/z (vep45K –panel G) Red dots represents levels of peak intensity for the spiked gargle/saliva samples, orange dots represents levels of peak intensity for COVID-19 RT-PCR negative control 1 samples, grey dots represent levels of peak intensity for COVID-19 PCR negative control 2 samples and blue dots represent levels of peak intensity for unscreened volunteer gargle/saliva samples. Blue dots with red circle borders are the values recorded for volunteer subject ID2

S1 peaks showed clear elevation in levels for the SARS-CoV-2 virus culture spiked samples, being on average 10^2^ x higher than the COVID 19 RT-PCR negative and volunteer gargle samples, where it was essentially not detected in 80% of samples. This marker alone could perform as a specific Corona virus MALDI ToF MS screening test with near 100% detection and specificity at 10^3^-10^4^ PFU SARSCoV-2 virions in a 10ml gargle/saliva sample.

S2b peaks may also be another key indicator as, although undetectable in the culture media spiked samples due to interference from albumin present in the media from FCS.

S2a peak intensity measures were more complicated. Although highly elevated in comparison to the control and random sample gargle/saliva group measurement was compromised as a peak of mass similar to that of S2b at 13,500 could be detected in a significant number of individuals. Many more peaks are seen in the lower mass range (below 20,000m/z) of saliva/gargle sample and probably arise from the abundant housekeeping proteins of residual bacterial flora. This “S2a like peak” of similar mass may characterize a specific bacterial species.

VEP protein peaks all showed elevated levels in the gargle/saliva spiked with SARS-CoV-2 culture media. However levels found in the control significantly overlapped, indicating these were not specific markers of COVID 19 but more generalised markers of virion membrane embedded proteins reflecting their common importance to enveloped viruses.

A notable sample from the volunteer group is that of sample ID2 which showed clearly elevated S1 and S2b, vep26–28k, vep34K, vep39–40K and vep45–47K and moderate elevated measurement of S2a.

## Immunological response and IgA levels

The local mucosal tissue barrier response to microbiological infection, including virus such as COVID-19, elicit secretion of IgA into the mucous and/or covering secretion such as tears and saliva (29). Peak masses matching that of Immunoglobulin light chains and the heavy chain of IgA were seen in the vast majority of gargle/saliva samples examined. Ig A and Ig G light chain are identical and that from IgM only slightly different on MALDI-ToF MS. However the heavy chain of IgG, Ig A and IgM are distinctly resolved on the MALDI-ToF mass spectrometer because of their different chain lengths (data not shown).

Only total Ig Light chains and IgA heavy chains were seen in the gargle/saliva samples. The intensity of these were measured in order to evaluate any correlation with other marker peaks (excluded in the analysis were the culture media virus containing “spiked” samples- as levels here were not physiological and reflected that of the base control gargle/saliva).

The levels of total Ig light chains were clearly differentiated between the two simultaneous COVID-19 RT-PCR negative control subjects. To note the higher levels seen in figure 6a were also reflected in S2a *like* (13,500m/z) protein peak levels for the same person (Figure 5). Within the subjects for whom COVID-PCR status had not been determined, the levels varied enormously from undetectable to the highest determined (Figure 6a). Subject ID2, the person with highest S1, S2b levels, had the third highest total Ig light chain levels.

**Figure 6.**
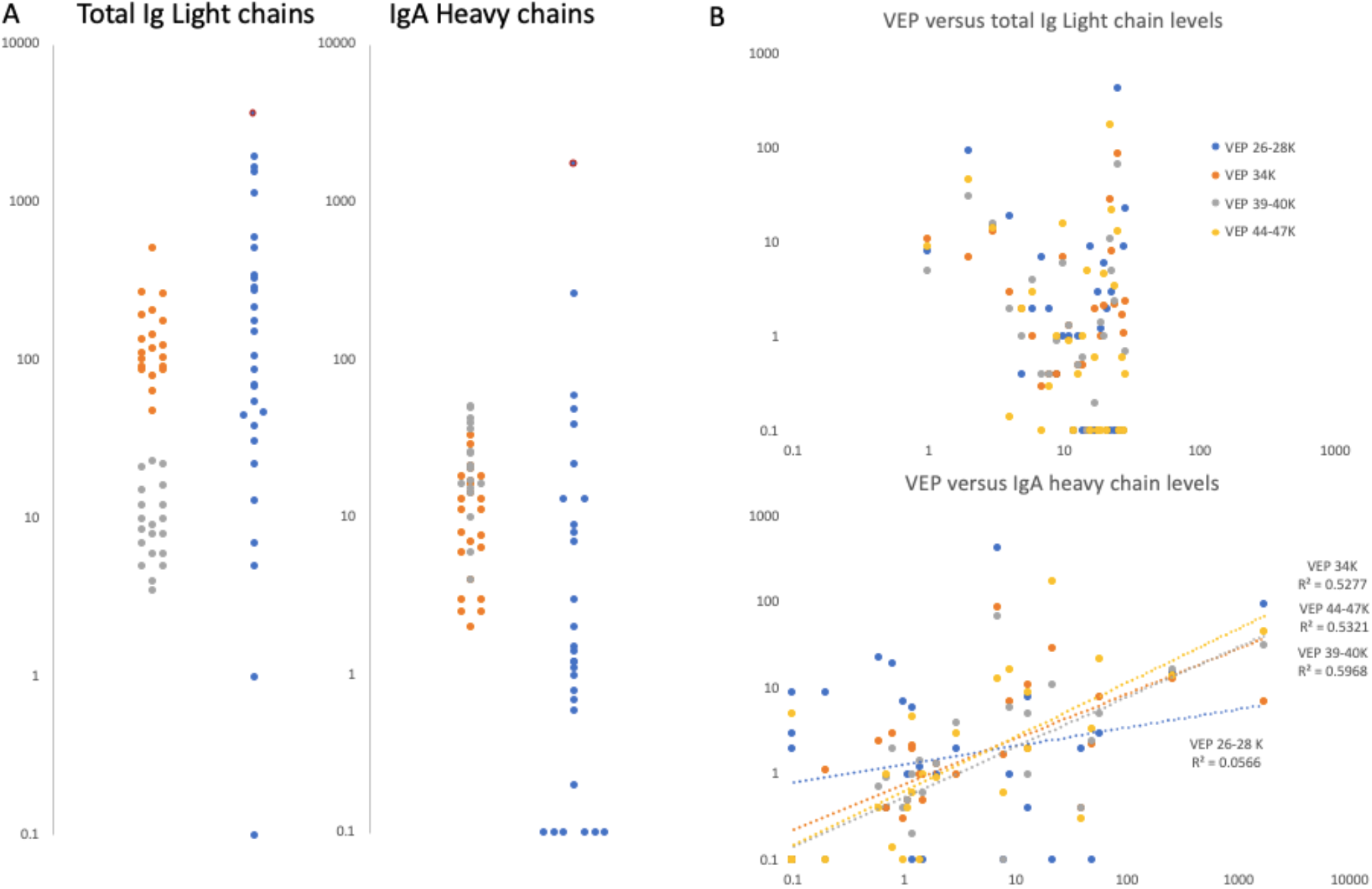
Gargle/saliva Immunoglobulin protein levels. Panel A are plots comparing the total Ig light chain and total IgA heavy chain peak intensity levels for two COVID-19 PCR negatives control individuals (grey and orange dots respectively) repeated 20 times each (n = 40), versus the same peaks intensity level of 30 volunteers (blue dots) who gave gargle/saliva samples but whose COVID-19 PCR status was unknown at the time. Blue dots bound by a red circle are the levels of total Ig light chains and IgA heavy chains found in volunteer ID2. Panel B are scatter plots of total Ig light chains (upper plot) and IgA heavy chain peak intensities (lower plot) versus VEPs peak intensities for the 30 volunteer gargle/saliva samples but whose COVID-19 PCR status was unknown at the time. No VEP level correlated with total Ig light chain level. Correlation were seen with Ig A heavy chain levels for vep39–40K (grey dots r^2^ = 0.60), vep44–47K (gold dots r^2^ = 0.53) and vep34k (orange dots r^2^ = 0.53) but not for vep26–28k (blue dots r^2^ = 0.06)

IgA heavy chain levels overlapped for samples from the two PCR-negative COVID-19 subjects and similarly, within the subjects for whom COVID-PCR status had not been determined the levels varied enormously from undetectable to the highest determined (figure 6a). The highest level being seen in the sample from the person with the highest S1, S2b levels, subject ID2.

It was noted that IgA heavy chains were only seen when elevated viral envelope proteins (VEPs) were detected in the mass spectra. To test a possible association we plotted total Ig light chain and IgA heavy chain levels against the levels measured for individual VEPs. Total Ig light chain levels did not correlate with the levels of VEPs detected; but IgA heavy chain levels did for vep34K, vep39–40k and vep45–47K (Figure 6b).

## Estimate of test costs

Adopting a MALDI-ToF MS based technology has had dramatic effects on reducing the costs in microbiological identification of infections and estimates have been as high as an 80% reduction overall [30]. A full evaluation of cost to achieve screening of millions in a population using this technique have to take into account discount on bulk orders. However, given that this system is generally reliant on extremely low volumes of generic reagents, the major cost is the purchase of the mass spectrometer. We have worked a basic model that mass spectrometry reagent analysis costs per test are less than $1 per sample. All the sample collection tubes and syringe filters associated in the process are more expensive at approx. $5 per sample at retail prices. Ignoring the latter peripheral costs of initial sample handling, we estimate that when including paying the cost of a mass spectrometer over 3 year in operation (at 100,000 samples a year per machine) the basic instrument-reagent analysis cost per sample is approximately $2.

## Discussion

Although simply applying the technique of MALDI-ToF mass spectrometry, as it has developed for identification of bacteria, for COVID-19 detection has been reported as promising [4], this approach is fraught with practical complications and analytical errors. Not least that the biochemistry focus that for bacterial identification the MALDI-ToF MS technique is optimized recognition of microbial pathogen hydrophilic, low to medium molecular weight, high abundance housekeeping proteins. SARS-CoV-2, an envelope virus, has little if any equivalent proteins and its virion contains a high abundance of medium to large membrane associated/embedded (predominately hydrophobic) glycoproteins. To develop and maximise a clinical efficient and cost effective MALDI-ToF mass spectrometry screening solution for COVID-19 and future viral pandemic screening, requires rational design based on an understanding of the chemistry, biology and physics of the pathogen-host interactions and also their unique marker biomolecules. This is further complicated by how such chemistry and proteins behave in MALDI-ToF mass spectrometry.

By targeting MALDI-ToF mass spectral detection to viral envelope glycoproteins a multidisciplinary team approach has achieved a multifaceted clinically MALDI ToF MS screening test for COVID-19 and future such pandemics that may arise. Further validation in relations to PCR detection in patients is needed but the system described promises a multifaceted diagnostic tool as follows:

1. Sensitive and specific identification of Corona virus in gargle/saliva by measurement of S1 and probably S2b protein peaks.
2. Together with S1 and S2b, very high level of a peak at 13,500m/z, presumed to be S2a, present in the *in vivo* sample is an additionally indicator of a corona virus infection (with high confidence);
3. More moderate elevation of S2a-like peak mass in the absence of elevated S1 and S2b peaks may be due to a similar protein/fragment arising from a microbiological infection of the oral respiratory tract, probably a bacterial housekeeping protein.
4. Elevated levels of VEPs are markers of viral infection
5. Elevated levels of total Ig Light chain is indicative of an oral upper respiratory tract infection
6. Elevated levels of gargle/saliva IgA heavy chain peak are indicative of viral infections

The final threshold for each component of the test are yet to be fully validated for computation software automation, but results are likely to be expressed as a heat map, the basic system being red, green and grey. The grey zone requiring retesting (see table 1).

**Figure 7:**
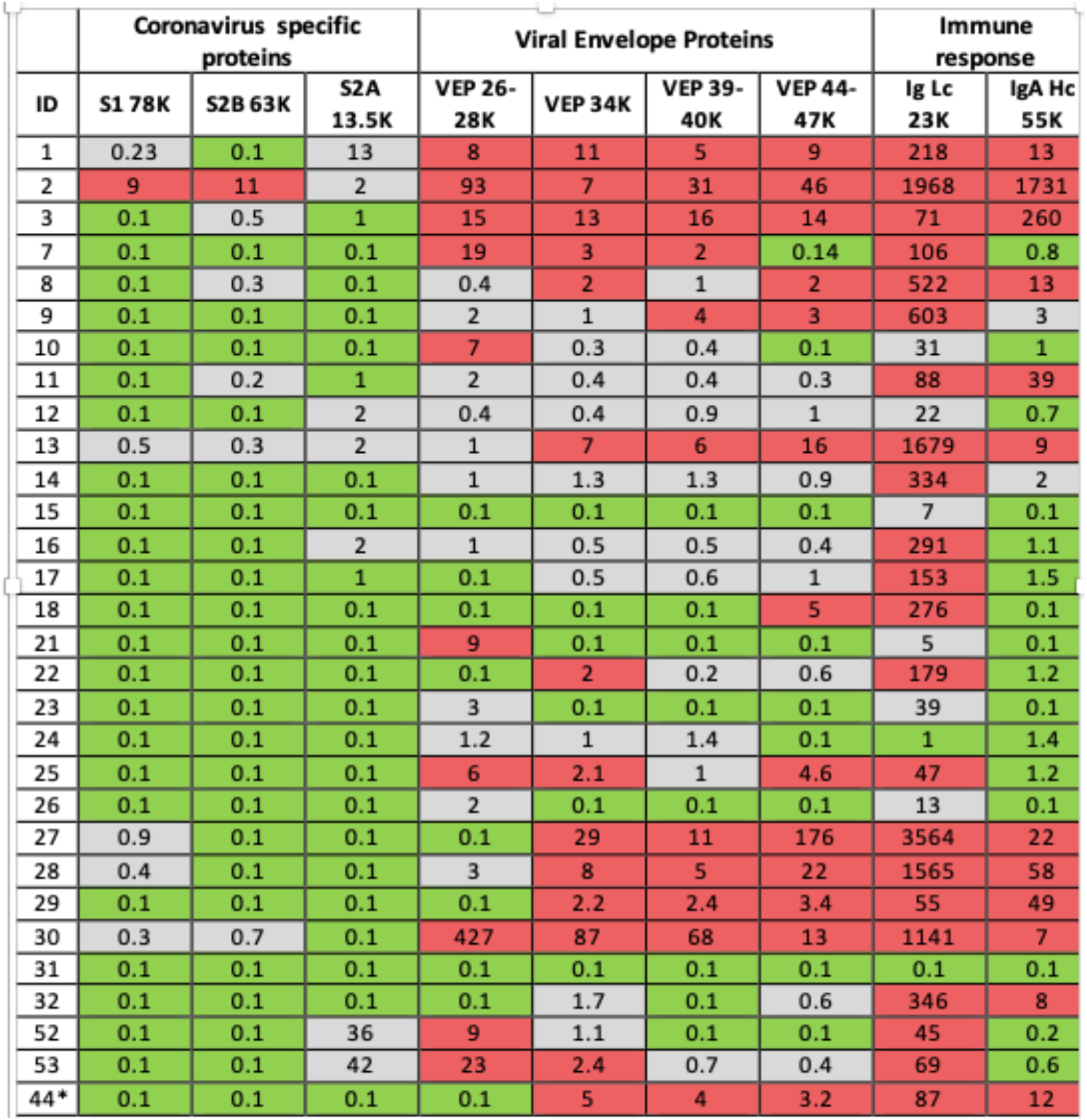
Illustration of clinical data output. Format is a heat map scoring of peak intensity relative to those values for SARS-2 corona virus spike protein fragments and viral envelope proteins, measured in gargle/saliva containing 1-10% culture media of SARS-CoV-2 virus at 10^4^ per ml PFU. Heat map associated with immunoglobulin intensities are based on the range found in the volunteers for which volunteer ID2 was identified as recovering from suspected COVID-19 infection. ID2 was repeated after 14days and his scores are shown under ID44*

In our preliminary study of COVID-19 RT-PCR untested volunteer samples for surveillance screening, volunteer subject ID2 stood out with elevated levels in all markers. In their 30s this individual worked in the music entertainment industry, they reported having been bed bound with Covid-19 symptoms, severe fatigue, loss of taste & smell and persistent cough in April (2020) 2 months prior to giving a gargle/saliva sample. At the time of sampling they reported to be 60–80% recovered from the symptom of fatigue which had persisted. In our follow-up they gave another sample 14days later and was also tested for COVID-19 by the NHS PCR service the same day. The analysis of the repeat sample showed no detectable corona virus S proteolytic fragment protein peaks; total Ig light chains and IgA heavy chain levels had fallen dramatically but were still elevated (see end of table 1 ID44*). The COVID-PCR result was reported as negative.

## Conclusion

Validation studies against saliva/gargle spiked with cultured virus are on-going and the current study indicate a close to 100% sensitivity if measuring S1 peaks alone as the indicator of Corona virus infection.

Direct comparison of the MALDI-ToF MS testing with RT-PCR detection of COVID-19 in clinical samples is needed. However, this MALDI-ToF MS technique cannot work on stored second sample Nasal-pharyngeal swabs as they will have either been heat inactivated (cooking the protein and viral envelope membrane) or stored in a transport media which contains SDS and or Triton which supresses ionization and similarly destroys the viral envelope.

In the absence of a freezer full of pre-collected 10ml gargle/saliva for which RT-PCR results are known, only a prospective study is possible. This is in process. We are working with several groups to collect gargle/saliva samples at presentation where a swab for PCR testing is being taken simultaneously. These groups will analyse the sample on their MALDI-ToF mass spectrometer instruments and with all the technical support and necessary key reagents provided.

As detailed, other markers measured in this technique may give further valuable clinical information such as other viral infections and magnitude of the mucosal humeral immune response.

## Data Availability

All raw spectral data (anonymised) is available on request

## Author Contributions

Conceptualization, RK IIes. and JL Heeney; methodology, RK Iles and JL Heeney, validation, JK Iles and R Zmuidinaite; mass spectral analysis, JK Iles and R Zmuidinaite; molecular constructs – in vitro investigations, G Carnell, A Sampson; data curation, RK Iles, JK Iles, R Zmuidinaite, G Carnell, A Sampson; writing—original draft preparation, RK Iles; writing—review and editing, JL Heeney, R Zmuidinaite, JK Iles, G Carnell, A Sampson; visualizations/images, RK. Iles, R Zmuidinaite, A Sampson; supervision, RK Iles and JL Heeney; funding acquisition, RK Iles and JL Heeney.

## Acknowledgments

The authors thank the willing volunteers who provided gargle saliva samples and in particular colleagues sharing pseudo virus and viral stains used in this study.

---

**Supplemental Figure s1:**
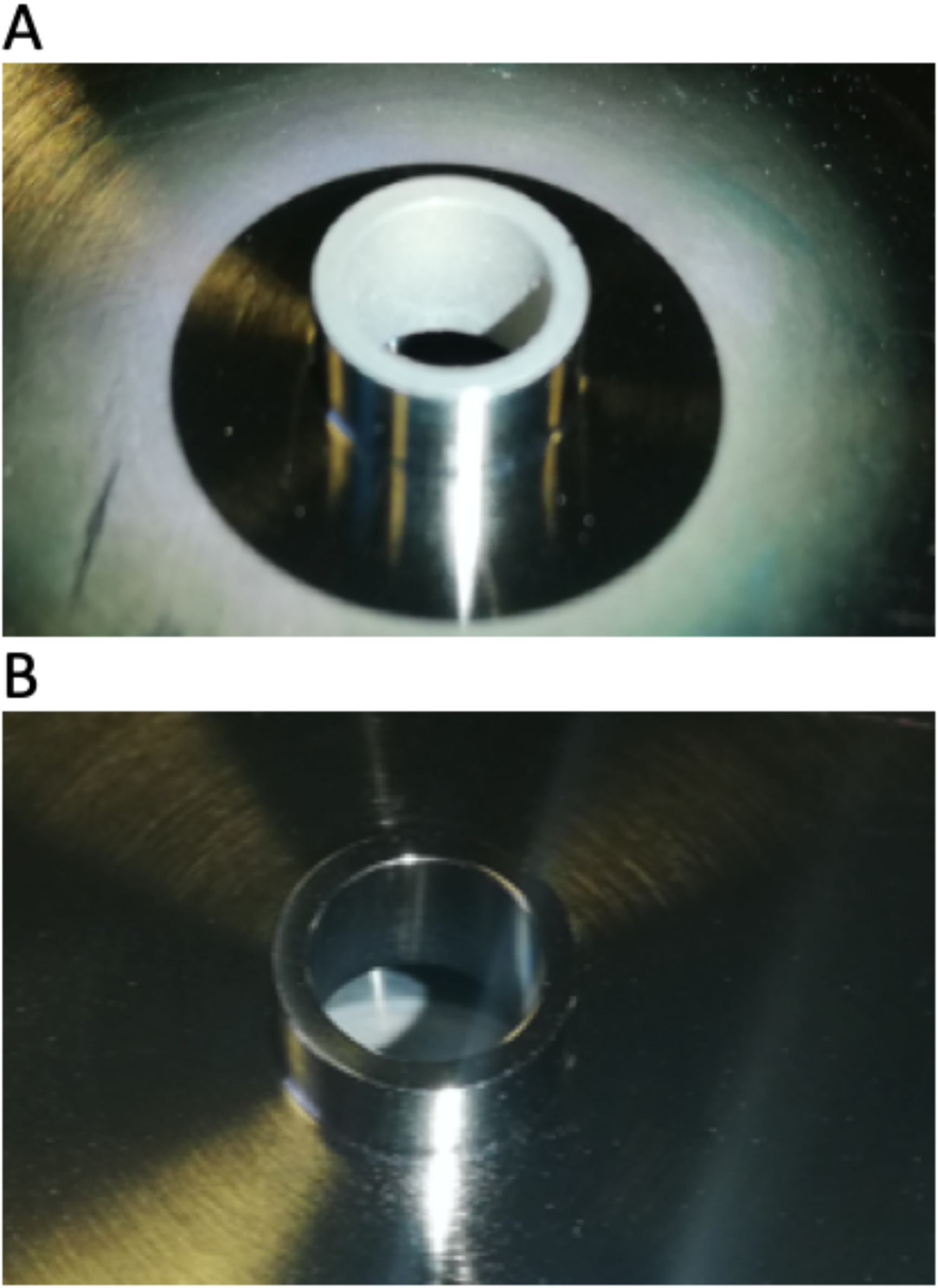
Fouling of primary lens from high throughput analysis of samples. Panel A shows the accumulation of material on the primary charge lens of the mass spectrometer as a result of matrix-reagent splatter after high throughput use in a simulated clinical setting. Panel B shows the cleaned optic.

